# Barriers to healthcare access: findings from a co-produced Long Covid case-finding study

**DOI:** 10.1101/2024.01.03.24300767

**Authors:** Donna Clutterbuck, Mel Ramasawmy, Marija Pantelic, Jasmine Hayer, Fauzia Begum, Mark Faghy, Nayab Nasir, Barry Causer, Melissa Heightman, Gail Allsopp, Dan Wootton, M Asad Khan, Claire Hastie, Monique Jackson, Clare Rayner, Darren Brown, Emily Parrett, Geraint Jones, Rowan Clarke, Sammie Mcfarland, Mark Gabbay, Amitava Banerjee, Nisreen A Alwan, the STIMULATE-ICP Consortium

## Abstract

**Background and aim:** Long Covid can be a stigmatising condition, particularly in people who are disadvantaged within society. This may prevent them seeking help and could lead to widening health inequalities. This co-produced study with a Community Advisory Board of people with Long Covid aimed to understand healthcare and wider barriers and stigma experienced by people with probable Long Covid.

**Methods:** An active case finding approach was employed to find adults with probable, but not yet clinically diagnosed, Long Covid in two localities in London (Camden and Merton) and Derbyshire, England. Interviews explored the barriers to care, and the stigma faced by participants and analysed thematically. This study forms part of the STIMULATE-ICP Collaboration.

**Findings:** Twenty-three interviews were completed. Participants reported limited awareness of what Long Covid is and the available pathways to management. There was considerable self-doubt among participants, sometimes reinforced by interactions with healthcare professionals. Participants questioned their deservedness of seeking healthcare support for their symptoms. Hesitancy to engage with healthcare services was motivated by fear of needing more investigation and concerns regarding judgement about ability to carry out caregiving responsibilities. It was also motivated by the complexity of the clinical presentation and fear of all symptoms being attributed to poor mental health. Participants also reported trying to avoid overburdening the health system. These difficulties were compounded by experiences of stigma and discrimination. The emerging themes reaffirmed a framework of epistemic injustice in relation to Long Covid, where creating, interpreting, and conveying knowledge has varied credibility based on the teller’s identity characteristics and/or the level of their interpretive resources.

**Conclusion:** We have developed recommendations based on the findings. These include early signposting to services, dedicating protected time to listen to people with Long Covid, providing a holistic approach in care pathways, and working to mitigate stigma. Regardless of the diagnosis, people experiencing new symptoms must be encouraged to seek timely medical help. Clear public health messaging is needed among communities already disadvantaged by epistemic injustice to raise awareness of Long Covid, and to share stories that encourage seeking care and to illustrate the adverse effects of stigma.

**Patient or Public Contribution:** This study was co-produced with a Community Advisory Board (CAB) made up of twenty-three members including healthcare professionals, people with lived experience of Long Covid and other stakeholders.

## Background

Long Covid is the patient-coined term for prolonged ill health following acute infection with SARS CoV-2.^1,2^ The World Health Organisation (WHO) defines post-COVID-19 condition (Long Covid) as occurring after ‘probable or confirmed SARS CoV-2 infection, usually 3 months from the onset of COVID-19 with symptoms and that last for at least 2 months and cannot be explained by an alternative diagnosis’,^1^ In March 2023, the Office for National Statistics (ONS) estimated that 1.9 million people were living with Long Covid in the United Kingdom (UK).^3^

Long Covid can cause varying symptoms and impact on daily life, including, but not limited to, fatigue, shortness of breath, headache, cognitive dysfunction, chest pain, muscle and joint pains, cough, disturbed sleep, and neuropsychiatric symptoms.^1,4–8^ These symptoms can have wide ranging negative social impacts that include inhibiting ability to engage in employment and creating financial burdens.^4,9–11^

Health inequalities exist between different population groups in England.^12,13^ Inequalities were evident prior to COVID-19 but the pandemic has exacerbated them and increased their visibility.^12^ COVID-19 resulted in higher infection and mortality rates for ethnic minorities and people living in more deprived areas in the earlier stages of the pandemic.^14,15^ Little is known about disadvantaged groups’ experiences of Long Covid but some evidence suggests that people living in more deprived communities and who belong to some ethnic minority backgrounds are more likely to experience Long Covid than those living in the least deprived areas or those from a White ethnic background,^16–18^ but conversely may be underrepresented in post-COVID services.^19^ This could suggest fewer people with Long Covid symptoms from these disadvantaged groups are receiving Long Covid care, which has the potential to increase health inequalities.

Stigma can occur with not conforming to standards that society calls normal.^20^ Stigmatised people are thus ‘disqualified from full social acceptance’ due to an attribute deemed to be ‘deeply discrediting’.^20^ Long Covid can be a stigmatising condition.^6,21,22^ Analysis of survey data of almost 900 people living with Long Covid in the UK found widespread stigmas associated with Long Covid.^21^ In this study, the Long Covid Stigma Scale (LCSS) measured three recognised mechanisms of stigma. These included direct experiences of discrimination (enacted stigma), expectations of poor treatment (anticipated stigma), and self-stereotyping based on negative connotations of having the condition (internalised stigma).^23–26^

A wide range of long-term conditions (LTCs) can be considered stigmatising^27^ and stigma is known to adversely affect help-seeking, as well as physical and mental health outcomes.^28^ Although anyone can experience stigma, its effects can be amplified for socioeconomically disadvantaged and ethnic minority groups.^28^ Experiences and expectations of discrimination can lead to poor engagement with healthcare services.^12^ It is therefore important to consider how multiple dimensions of disadvantage intersect when exploring health stigma and other barriers to care.

We aimed to co-produce with people with lived experience of Long Covid a community-based approach for identifying people with probable Long Covid, who have not yet received a formal clinical diagnosis, for the purpose of understanding the barriers and stigma they face when attempting to access support and care.

## Methods

This work forms part of the National Institute for Health and Care Research (NIHR) STIMULATE-ICP (Symptoms, Trajectory, Inequalities and Management: Understanding long COVID to Address and Transform Existing Integrated Care Pathways) study. Methods were detailed in a published protocol.^29^

### Co-production with a Community Advisory Board (CAB)

Research co-production is where researchers work with public contributors, including patients, to design and/or conduct research.^30,31^ A Key benefit is generating knowledge that is more meaningful to the population group being studied.^31,32^ In this study, a Community Advisory Board (CAB) was formed. This was made up of ten health care professionals (including seven who are also living with Long Covid), eight people living with or caring for children with Long Covid and five other stakeholders from voluntary organisations and local public health bodies.

Six CAB meetings took place between February 2022 and July 2023. Discussions in these meetings centred around defining and refining study aims and methodology, planning the implementation of study findings and dissemination. The CAB also contributed to the design, review, and production of study documents, and members are co-authors on this paper.

### Study design

The purpose of using an active case finding approach was to find individual cases of probable Long Covid that have not been clinically diagnosed. This study was initially conducted in the London Borough of Camden, England. Two other sites later joined the study: the London Borough of Merton, and the county of Derbyshire, England. Health inequalities exist within each of these localities.^33–35^ Camden is the most ethnically diverse area – 41% of people who reside in it come from an ethnic minority background.^33^ It is estimated that 37% of residents in Merton are from an ethnic minority group and inequalities exist between the east and west of the borough.^34,36^ In contrast to the London sites, a large part of Derbyshire is rural and 6.3% of the population are from an ethnic minority background.^37^

A leaflet raising awareness of the study and some of the most common symptoms of Long Covid was disseminated within communities in the participating localities, with the aid of community organisations, local authorities and social prescribing teams.^29^ Individuals who met the eligibility criteria were assumed to have probable Long Covid and were invited to take part in a qualitative interview. The eligibility criteria were agreed with the CAB (figure 1, supplementary material A and B) and are detailed in the study protocol.^29^

**Figure 1.**
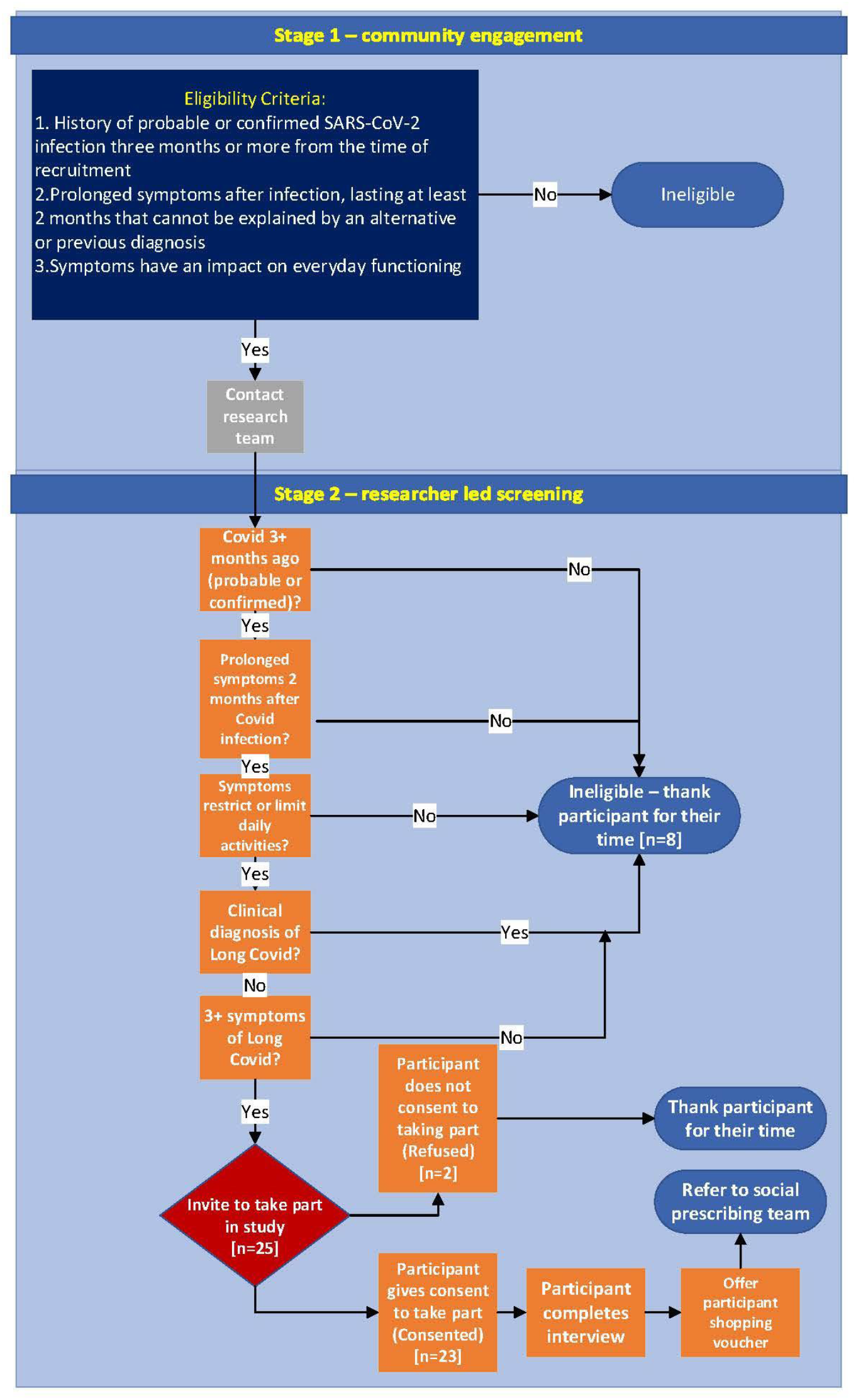
Study eligibility flow chart.

### Qualitative research data collection

Semi-structured interviews were completed with people with probable Long Covid across Camden, Merton and Derbyshire. The interview topic guide was co-produced with the CAB. This guide consisted of predominantly open questions investigating barriers to obtaining care for Long Covid symptoms. It also included some questions from the LCSS^21^ exploring possible stigmatising experiences across the three mechanisms of enacted (overt experiences of being treated unfairly), anticipated (expectation of being treated unfairly) and internalised (adopting negative stereotyping and applying to self) stigma. Prompts were used to find out the context of these experiences.

All interviews were conducted by DC between August 2022 and May 2023, most completed by videocall. Four were completed via telephone call. Informed consent was obtained from each participant prior to undertaking the interviews. Participants were reminded at the beginning of the interviews that being eligible for the study did not constitute a clinical diagnosis of Long Covid. Upon completion of the interview, participants were offered a shopping voucher as a thank you gesture for their time, and a referral to a social prescribing service to receive personalised support. Social prescribing aims to improve mental and physical health through referral to community-based services for holistic support.^38^ Participants were also offered a letter indicating participation in the study that could be taken to their primary care team. A full outline of the methods is in the study protocol.^29^

### Analysis

Thematic analysis, based on the approach described by Braun and Clarke.^39,40^ was employed to analyse the interviews. After becoming familiar with the transcripts, DC completed initial coding using predominantly an inductive approach using NVivo. Some codes matched the concepts studied in the LCSS so more deductive coding was applied in relation to these questions. MR completed additional coding on a sub-sample of transcripts to ensure intercoder reliability. Once coding had been completed the DC, MR and NAA met multiple times to discuss potential themes. The LCSS informed the creation of themes related to experiences of enacted, anticipated, and internalised stigma.^21^ A thematic framework was developed in light of the findings and agreed upon before drafting this paper.

## Results

Twenty-three semi-structured interviews were completed with participants with varying symptoms consistent with Long Covid, who reported not having a known clinical diagnosis. Information regarding these symptoms can be found in supplementary material C. Participants’ characteristics can be found in Table 1. The majority of participants were from the Camden locality. Sharing study materials via voluntary organisations was the most fruitful way to access communities in this study.

**Table 1:** Participant Characteristics.

|  |  |
| --- | --- |
| **Gender** |  |
| Female | 16 |
| Male | 7 |
| **Ethnicity** |  |
| White British | 8 |
| White/White other | 6 |
| Asian/Asian British | 5 |
| Black/ Black African/Black | 2 |
| Caribbean/Black British |  |
| Mixed/Multiple ethnic groups/other | 2 |
| **Age** |  |
| 30-39 | 5 |
| 40-49 | 5 |
| 50-59 | 6 |
| 60-69 | 4 |
| 70-79 | 3 |
| **Index of Multiple Deprivation (IMD)** |  |
| **quintile \*** |  |
| 1 = most deprived areas | 4 |
| 2 | 5 |
| 3 | 7 |
| 4 | 3 |
| 5 = least deprived areas | 3 |
| Unknown | 1 |
| **Disability status** |  |
| Disabled | 6 |
| Not disabled | 17 |
| **Long Term Conditions (LTCs) present** |  |
| **before COVID-19** |  |
| Yes | 19 |
| No | 4 |

\* *Index of Multiple Deprivation which measures areas of deprivation in localities within England. English indices of deprivation 2019. Available from: https://www.gov.uk/government/statistics/english-indices-of-deprivation-2019.*

A minority were unsure about their Long Covid diagnosis status. It was assumed that these participants did not have a formal diagnosis and so they were included in this study. This was on the basis that some of these participants noted that Long Covid had been suggested as a possibility by healthcare professionals (HCPs) but they had had no follow up investigations. Two of the participants believed that referrals might have been made but these had not been fulfilled at the time of interview.

Four key themes were identified during analysis: 1. Uncertainty and Long Covid awareness 2. Barriers to engagement with the healthcare system 3. Experiences and perceptions of stigma and discrimination and 4. Sources of support. The following sections focus on these themes.

### Theme 1: Uncertainty and Long Covid awareness

#### Awareness of Long Covid and Long Covid support services

A lack of adequate awareness of Long Covid was prominent in the accounts of participants. Some participants were not aware of the existence of Long Covid as a health condition, others had some awareness but did not equate their symptoms to Long Covid.

> ‘My symptoms as such have never really gone away but I’ve not always attributed them to Long Covid or whatever’. (female, aged 60-69, White British)

Participants were more likely to describe not feeling ‘normal’ or not feeling ‘the same since’ their COVID-19 infection. Others tended to connect Long Covid with more ‘severe’ symptoms. Two participants expressed how they only became aware of Long Covid upon engagement with the study information.

> ‘I wasn’t aware of it, but now you’ve kind of made me aware’. (female, aged 30-39, Bangladeshi-British)

It was only through engagement with the study leaflet that the possibility of Long Covid was considered. Other participants suggested that the study information helped to reinforce their belief that their symptoms were caused by Long Covid.

Some participants assumed lack of knowledge and awareness of Long Covid within the National Health Service (NHS).

> ‘They still don’t know what to do and they don’t know what to say’. (female, aged 70-79, White British)

There was a lack of awareness of how to obtain care for symptoms and some felt there was no support available.

> ‘I don’t think there’s any cure. There’s nothing they can do’. (female, aged 70-79, Jewish)

Furthermore, some participants indicated that they would seek help if they felt that treatment was available. A need for increased awareness, both within the community and in healthcare, was stressed by some.

> ‘Awareness is so important in order to even get a diagnosis or think, well, it could potentially be that, and it’s the same with professionals’. (female, aged 30-39, Bengali)

#### Self-doubt

Self-doubt prevented some people with Long Covid symptoms from seeking help. Participants were sometimes unsure if their symptoms were caused by Long Covid, and some explained their symptoms as potentially resulting from ageing, as being related to menopause, or being stress-related.

> ‘Because I’ve had just a lot of life changes, you know, like in terms of my career, what’s going on with work, with stress, all of that, I’ve then as a result not really felt that comfortable saying this might be something else as well [laughing]’. (male, aged 30-39, Black British Caribbean)

There was also evidence of participants downplaying symptoms or comparing their experiences to others.

> ‘I didn’t really think it would be right to say it was Long Covid is having read about people suffering from Long Covid as having really, really severe symptoms and not being able to do anything and thinking this doesn’t apply to me’. (male, aged 60-69, Asian British)

Sometimes, interactions with HCPs reinforced this self-doubt. For example, women who suspected that their symptoms could be menopause-related had this confirmed by GPs without further investigation.

> ‘I kind of spoke to a doctor about the fatigue. And the dizziness and stuff. And he just went, oh, yes, it just sounds like perimenopause, just got to get on with it and that was kind of it. So, that was as far as it went because I thought I don’t really want to bother them again’. (female, aged 40-49, White)

Likewise, if Long Covid was not suspected by HCPs, some participants rejected it as a possibility.

> ‘So I started Googling it, and then I thought, This sounds like what I have. But because the GPs didn’t mention it, I just thought, they’re the professionals. They know me best in terms of medically, and they know my medical history. So I was leaning towards oh, it can’t be’. (female, aged 30-39, Bengali)

Participants trusted HCPs.

> ‘You think you’re talking to a consultant and health professionals that they know better’. (female, aged 50-59, White)

### Theme 2: Barriers to engagement with healthcare

#### Hesitancy around seeking support from the healthcare system

Hesitancy around seeking treatment or care was apparent for different reasons. One participant was concerned that if his symptoms were not associated with Long Covid, further investigation might be needed.

> ‘There’s a possibility that it’s not Long Covid then I’d have to… Like that would involve working out what is going on’. (male, aged 30-39, Black British Caribbean)

Similarly, another participant expressed that he did not want to be prescribed medication and this was preventing him from reporting symptoms to a HCP.

Another participant explained that she was worried about engaging with healthcare in case assumptions about her ability to fulfil her parenting responsibilities were made.

> ‘Sometimes I am afraid if I say to them that, you know, they feel that I am not capable to look after my son and my kids, you know. It’s kind of a worry behind my head. I wouldn’t like to mention it.’ (female, aged 40-49, Bangladeshi)

Reluctance to engage was sometimes tied to not wanting to burden the NHS.

> ‘At the moment, I’d probably feel a bit like I was wasting their time… Because I just feel like at the moment the NHS is in a bit of a state’. (male, aged 30-39, White)

Participants felt that their symptoms were not a priority to a health system which they considered to be overloaded.

Additionally, engaging with healthcare was considered a part of a non-help seeking identity for some participants. Some participants expressed how they would have to be exceptionally unwell to consider reaching out.

> ‘I’m not that kind of, I don’t think I can even knock someone’s door to ask for help’. (female, aged 50-59, Mixed Race)

Hesitancy in accessing care was felt to be exacerbated by perceived problems in primary care, such as long waiting times for appointments, limited appointment times, only being able to focus on one symptom during primary care consultations or waiting for secondary care referrals for individual symptoms to be fulfilled before any further steps are taken.

#### Nature of multi-system symptoms makes accessing care difficult

For some, the very nature of Long Covid symptoms made accessing treatment or care difficult. One participant was hesitant to seek care from her GP because of attribution of new symptoms to pre-existing health problems.

> ‘They tend to always assume that my symptoms are consistent with my chronic long-term health problem.’ (female, aged 50-59, White British)

The fluctuating and relapsing nature of symptoms, which can occur with Long Covid,^1,4–6^ acted as a barrier to obtaining care.

> ‘Just when you think, oh, they’ve gone, up they pop’. (female, aged 50-59, White British)

Sometimes this meant that symptoms were not necessarily present at the time of the healthcare consultations or would disappear soon after. Thus, the changing nature of symptoms can mean that patients do not return for follow-up care.

Sometimes one symptom was more prominent than other experienced symptoms. Unsurprisingly, these participants sought care for the symptom impacting them the most, which resulted in other symptoms being overlooked by the participant and HCPs. This can result in the pattern of symptoms of Long Covid being missed.

One participant had a flare up of her existing health conditions after an acute infection with COVID-19, despite this being stable for a long period of time, and so her other symptoms were neglected:

> ‘I’ve seen my GP, but that’s been it to be honest. I think once all of my stomach issues kind of kicked in and, you know, it was a bit of a kind of panic [laughs] because you know it can be kind of quite hard and fast when it comes on and so the focus really, probably went on to that to be honest’. (female, aged 40-49, White British)

Some participants had experiences of symptoms being attributed to mental health conditions by HCPs including one who was offered anti-depressant medication. She acknowledged that she had been experiencing low mood, but this was because of her concerns around not being listened to.

> ‘I feel upset because he tried to give me Prozac for it, and I’m trying to explain, it’s not a Prozac situation here’. (female, aged 50-59, White British)

### Theme 3: Experiences and perceptions of stigma and discrimination

#### Experiences of discrimination

A range of perceived experiences of discrimination, both within healthcare and in the community, were evident from the interviews. Participants reported compound experiences of disadvantage shaped by the intersection of their identity dimensions with experiencing Long Covid leading to layered stigma.

A few participants described ageism.

> ‘I always feel that your age is put against you’. (female, aged 50-59, White)

Gender discrimination was implicated in the interviews of some female participants, and this was in the form of GPs lacking knowledge of how to treat women, symptoms being more likely to be attributed to mental health and feeling belittled by HCPs, as one participant described in her interaction with a male consultant:

> ‘I did find he was very condescending and patronising and, in my view, I felt that was because I was a woman’. (female, aged 40-49, White British)

Symptoms and participant knowledge of their own bodies were also reported to be dismissed by HCPs.

> ‘He literally dismissed it, that was it. And I think that was the problem with the doctors, they just dismiss things sometimes if they don’t want to deal with it. So, he literally dismissed it. There was no, why is this happening. What can we look in to, you know, is it COVID, is it this. There was nothing’. (female, aged 40-49, White)

Feelings surrounding a lack of understanding and support were prominent among the participants. A few participants talked about concerns from others regarding transmissibility, not experiencing empathy or sympathy, not being included in social activities, being judged, being ignored, being perceived as ‘lazy’, or receiving unsolicited suggestions for symptom relief which sometimes carried undertones of blame for not doing enough to keep well.

One participant revealed how she only socialised with people with LTCs because of the lack of understanding from others without chronic illness.

> ‘My contacts tend to have come with other people who are ill because it’s very difficult to have social interaction with people who aren’t ill because they understandably lose patience if you can’t physically meet up with them’. (female, aged 50-59, White British)

Additionally, a few participants talked about a lack of understanding and support from employers and colleagues. This was usually evident in comments around inefficiency or questioning symptoms and sick leave:

> ‘My friend and colleague said, “Oh, you are trying to make excuses to take annual leave.”… and it made me feel a bit like, you know, upset. And I said to the person, “You know, I wasn’t making it up. I was unable to get out of bed and I was really struggling to breath[e] some days”’. (female, aged 40-49, Bangladeshi)

#### Anticipated stigma

Some participants expected discrimination to occur because of their Long Covid symptoms. Sometimes, this was due to past stigmatising experiences. For some participants, having a history of being dismissed prior to Long Covid impacted their view of help-seeking. For one participant this dismissal had previously occurred due to her weight.

> ‘it’s almost as if they bring the weight to the focus and then I feel guilty for taking up time’. (female, aged 60-69, White British)

One participant expressed worries surrounding engagement without proof of his symptoms.

> ‘Unless there’s something concrete that I could say, look this is, there is a physical symptom, I’m not making it up, I don’t really want to engage [laughing]… I do worry that if it is dismissed that then…they could put a little thing down that I’m a bit of a hypochondriac. So I’d rather not bother’. (male, aged 30-39, Black British Caribbean)

Others similarly suggested that they did not want to be labelled an ‘idiot’, receive a ‘quizzical look’ or be considered ‘awkward’. This participant also described how he previously contracted Monkeypox, sought support and was ‘rebuffed a few times’, despite experiencing symptoms and identifying as being in the at-risk demographic, before being given a positive diagnosis weeks later. For these participants, their avoidance of dismissal was preventing them from seeking care for their symptoms.

Disbelief was seen as resulting from a lack of understanding as well as due to varying symptoms. Some participants felt that this disbelief extended beyond the community into the medical profession. One participant asked the interviewer *‘how accepting is the health care profession that Long Covid is real?’* (female, aged 60-69, White British), suggesting that she was not sure if Long Covid is taken seriously by HCPs.

There were participants who had interactions with others who expressed disbelief, but mostly this was a perception, and some participants saw dismissal as proof of disbelief.

Participants feared judgement from others due to the impact of symptoms or other people’s presumptions of the impact of symptoms.

> ‘They will presume that…you’re not on the ball’. (female, aged 60-69, White British)

This fear of judgement was a common reason for participants not talking to others about their symptoms. One participant, early on during her interview, stated that she did not feel comfortable talking about mental health with her GP.

> ‘I don’t feel that comfortable to say everything about it [mental health] and feel the questions sometimes the doctor asks like I would not be very happy. Actually not that comfortable. Maybe I am from a community with religion and some of the questions [answers] I am not that comfortable to share’. (female, aged 40-49, Bangladeshi)

Later, she suggested that she would be apprehensive about sharing details of her symptoms with a potential partner as they may not want to marry someone with a LTC. For some participants, the interview was the first time they had talked about their symptoms and/or the possibility of symptoms being related to Long Covid.

Employment worries were evident throughout the interviews. One participant had to switch from a more physically demanding role to a ‘desk job’ because of his symptoms. Participants often felt they were not performing as well at work as they did before.

> ‘A couple of times I’ve had to say I need to go home because of this…I feel like I’m taking the Mick [not taking work seriously] myself’. (male, aged 50-59, Asian)

Another participant (male, aged 60-69, White) suggested that if he was looking for a new role, he *‘might keep it [Long Covid] to [himself]’* to avoid negative views preventing him from finding employment.

Another suggested that as Long Covid is not considered a disability, you would not have employment protections.

> ‘It’s not considered a disability… you could just be fired just like, you’re not performing. And you go, well, I’ve got Long Covid. They go… we’re not legally obliged to consider that so, tough’. (male, aged 30-39, White)

This perhaps goes some way in explaining the worry surrounding employment performance.

#### Internalised stigma

There were indications of shame from participants about their illness and associated symptoms. When participants were asked if they felt embarrassed about their illness or physical limitations, their answers focused on their symptoms and not performing as well at work and socially. There were a few participants who felt embarrassed as they were not as fit, ‘motivated’ or ‘physically strong’ as they felt they should be.

However, some were embarrassed by illness in general. One participant explained this as resulting from confusion surrounding being ‘infectious’ and another felt that their symptoms were so ‘vague’ it could lead to disbelief from others. One participant suggested that if others know you are ill, *‘it changes their perception of you’.* (female, 40-49, White British)

Symptoms and not being able to do the same things as before were the main reasons why participants felt of less value due to Long Covid.

> ‘It kind of affects me, like across the board with the tiredness. And the forgetfulness and stuff. You just feel like a lesser person’. (female, aged 40-49, White)

Participants stated they felt different to others because of their symptoms, and some participants compared themselves to others who seemed to recover from COVID-19 more quickly than them.

When asked if anything has happened to make them feel embarrassed, tainted or different, participants often stated that this was an individual thing and that others have not contributed towards these feelings*: ‘No, I just think it-Again it’s a personal thing that I feel sometimes’.* (female, aged 40-49, White British)

### Theme 4: Sources of support

#### Sources of treatment outside of the NHS

Just under half of the participants accessed treatment, care, and support outside of the NHS. This included using alternative therapies and remedies, experimenting with vitamins and supplements, consulting with family and friends who are trained HCPs, resorting to private health care, as well as making behavioural changes through diet and exercise. One participant asked a HCP during a secondary care appointment to prescribe her medication after her GP initially refused.

> ’It was my last port of call, I said, “I’m desperate to get this, if I have it privately it’ll cost me a lot.” “What is it?” he said, and then he looked it up and he said, “I have heard about it, people have talked to me about it, but I’m not sure if it will work. It may be one of those solutions that are a bit cranky but I’ll let you try.” And he prescribed it’. (female, aged 70-79, White British)

Some participants felt they must be creative when seeking out care and support as they perceive mainstream healthcare to be inadequate.

#### Social and community support

Social and community support such as peer support, gym/physical and outdoor activity groups, as well as pharmacies for advice-giving were considered useful for people living with Long Covid. Three participants used social media Long Covid support groups to seek advice regarding their symptoms. Additionally, most participants were receptive to the social prescribing referral offered by the study.

Support that included social interaction seemed most popular amongst participants.

> ‘It’s a bit like anything isn’t it. If you’ve experienced something yourself then it’s nice to know that it’s not just you that it’s happening to and that you’ve got someone to relate to’. (female, aged 50-59, White)

Participants described sharing their experiences within their social circles because*: ‘a lot of people are saying they’ve got the same thing’* (female, aged 70-79, Jewish). One revealed that she had shared the study information with her sisters because *‘they’re having the same problem’* (female, aged 30-39, Bangladeshi-British), referring to her Long Covid symptoms.

Financial support was mentioned in relation to recouping funds that had been spent on alternative therapies or supplements, as well as for those who are unable to work fulltime. Similarly, employment support was considered beneficial for people living with Long Covid. Some expressed how hybrid working and working flexibly had been advantageous to them when suffering Long Covid symptoms.

#### Desired healthcare expectations

Participants offered ways to decrease barriers to healthcare for those living with Long Covid symptoms. Changes to the primary care system such as making it easier to get an appointment, more face-to-face appointments, longer appointment times that are*: ‘more understand[ing] of the needs of the patient’* (female, aged 40-49, Bangladeshi) were offered as solutions to meet the needs of those with Long Covid.

Participants wanted to feel listened to, unpressured by time constraints, have room to explore symptoms, reassurance that they are not wasting time, and to receive advice, treatment, signposting or referrals where appropriate from HCPs.

> ‘You would want to feel that they had the time to consider what you’re saying and what our symptoms are and not feel pressurised by lack of time’. (female, aged 50-59, White British)

Dedicated services for Long Covid were popular among participants, and these were in the form of helplines, webpages, specific appointments in GP practices for Long Covid and specialist clinics for people with Long Covid symptoms.

> ‘If there was a body that you could go to where it would be, you’d be asked questions specifically about your experience with COVID and your experiences now and your symptoms now. Perhaps if there was a dedicated place you could go to for that, that would make it a lot, because then you’d be, people would be specifying on that subject rather than thinking oh it could be HRT or it could be this or could be that. At least they’d be asking you about that specifically at first’. (female, aged 60-69, White British)

Some participants felt that there should be ways to access these services without visiting a GP.

## Discussion

This study developed a community-based approach to identify and include participants with probable Long Covid. The co-production process meant the inclusion criteria directly reflected the experiences of people living with Long Covid, which in turn meant that the study leaflet resonated with the intended audience. Subsequently, this led to engagement from people living with Long Covid symptoms that had previously not come forward for support.

It is evident from this analysis that a lack of adequate awareness of Long Covid, self-doubt, hesitancy in seeking care and the nature of multi-system symptoms act as barriers to seeking care for Long Covid symptoms. Participants’ interviews also revealed experiences of perceived discrimination, and anticipated as well as internalised stigma and which can further impede help-seeking behaviour.

Stigma leads to the discounting of the experiences of people with Long Covid due to negative stereotyping. When multiple forms of marginalization are compounded by experiencing a stigmatised health condition, this results in layered or intersectional stigma,^41,42^ which has been described in this study.

Some findings from this study show similarities with other Long Covid studies. Most notably, the varying range of symptoms experienced,^4–8^ the lack of Long Covid awareness among some population groups,^7,43,44^ self-doubt regarding Long Covid as a cause for symptoms,^7^ symptoms as a barrier to accessing care,^43^ problems accessing primary care,^6,10,45^ experiences of symptoms being dismissed,^10,46,47^ seeking support outside the NHS and making behavioural changes,^6,11,45–48^ as well as stigma^6,47,48^ and discrimination^45^ which can all act as barriers to obtaining support, care and treatment for symptoms.

Parallels can also be drawn with Ireson et al.^46^ who suggest the experiences of people living with Long Covid, equate to epistemic injustice. Epistemic injustice is inequality surrounding knowledge^49^ and can occur as ‘hermeneutical injustice’ (where an individual does not have the means to interpret their ‘experiences’) and ‘testimonial injustice’ (discriminatory mistrust of an individual’s knowledge).^49^ As Bhakuni and Abimbola explain, testimonial injustice happens when a hearer prejudicially ascribes lower credibility to a speaker’s words, for example, through acts that silence, undervalue, or distort. Hermeneutical (or Interpretive) injustice happens with someone struggles to make sense of and share their experience of the world, owing to a gap in available legitimised collective sensemaking resources. This particularly occurs when the experiences of marginalised individuals or groups are not understood by themselves or by others because those experiences do not fit any concepts known to them (or to others).^50^ Our findings fit under both frameworks (Figure 2).

**Figure 2.**
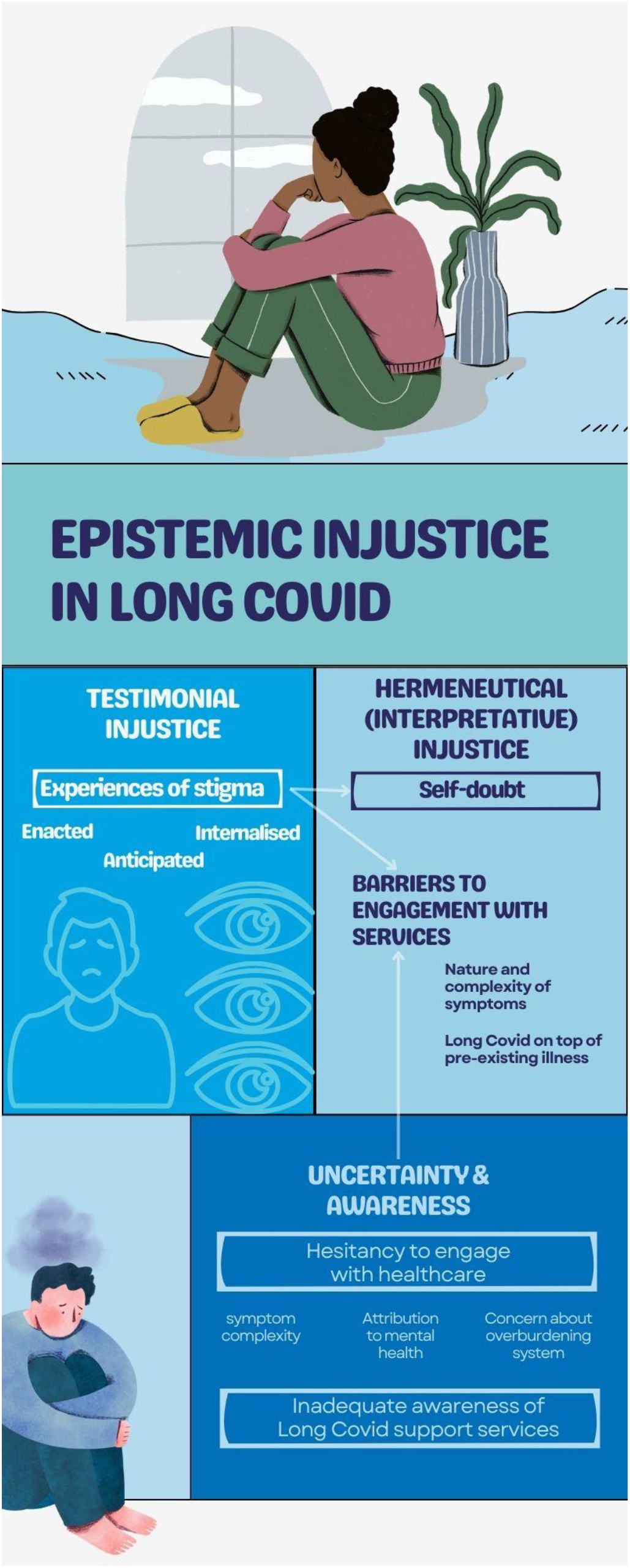
Epistemic Injustice in Long Covid as a framework for the study findings.

Epistemic injustice within health systems^51^ and in relation to other chronic conditions has been studied.^52^ There is inherently a power imbalance in the patient-HCP relationship. HCPs are unsurprisingly considered more medically knowledgeable, and they also have the power to decide suitable treatment options for different patients. These options are decided based on which patient narratives HCPs define as believable or unbelievable (testimonial injustice).^51,52^ This can result in some patients being dismissed, as in this study, and in similar under-recognised and under-researched conditions such as chronic pain or myalgic encephalomyelitis. ^52,53^ On the other hand, patients may not have the ability to convey their symptoms in a way that is understandable by HCPs (hermeneutical injustice).^51,52^ These experiences are compounded by inadequate awareness, knowledge and understanding of Long Covid, both within the community and at the healthcare level, due to its novel status, and also perhaps due to inadequate reference to Long Covid as a result of deficient public health messaging about the potential long-term health effects of COVID-19.

Experiences of anticipated stigma were presented in this study. Choices about revealing illness status are often bound up with expected stigmatising consequences for people with chronic conditions.^54^ Others have also confirmed that mental health stigma can prevent help-seeking^28,55^ and the intersection with ethnicity, culture and gender can increase its impact.^28^ Additionally, experiences or feelings of social exclusion are not uncommon in health stigma literature^28^ and weight-based prejudice as a reason for avoiding healthcare has been highlighted by others too.^28,56^

Similarly, internalised stigma, including feelings of shame or making self-comparisons with others, has been associated with other chronic conditions.^54^ One explanation could be because individuals have internalised expectations of how they should be, and if they do not meet these expectations, they feel stigmatised.^27^ For people with Long Covid, this could partly be due to COVID-19 being described as a ‘mild’, self-limiting illness that most people recover from quickly – a definition which is evidently contested.^46,47,57,58^ Those who experience ongoing symptoms from COVID-19 are situated outside this dominant definition of ‘mildness’ which can result in feelings of being flawed.^27^

Experiences of stigma can prevent people living with Long Covid from seeking and/or receiving much-needed support for their symptoms. They can lead to self-doubt and hesitancy in seeking care. Self-doubt surrounding the cause of, and finding other explanations for, symptoms can act as a barrier to obtaining care for chronic conditions.^54^ Bury explains this as a way of attempting to regain some ‘control’ as well as minimise the perceived damage to identities.^54^

Unlike in other studies,^6,47,48^ these participants did not identify online communities as a key source of support. This could be due to the lack of awareness of Long Covid in this group. Participants were receptive to potential forms of social and community support, nonetheless. Stigma can be minimised when people know that they are not alone in their experiences. Additionally, financial and economic support were considered useful, strengthening the argument that Long Covid, like other LTCs, can create social and economic burdens.^4,9^

The study recommendations are summarised in figure 3. We are working on producing an online tool to help facilitate help-seeking in people with Long Covid who are struggling to access support. It is important to recognise that although all participants in this study reported symptoms consistent with Long Covid, alternative diagnoses are possible. Regardless of the diagnosis, people living with these symptoms need to be encouraged to seek medical help to get the support that they need.

**Figure 3.**
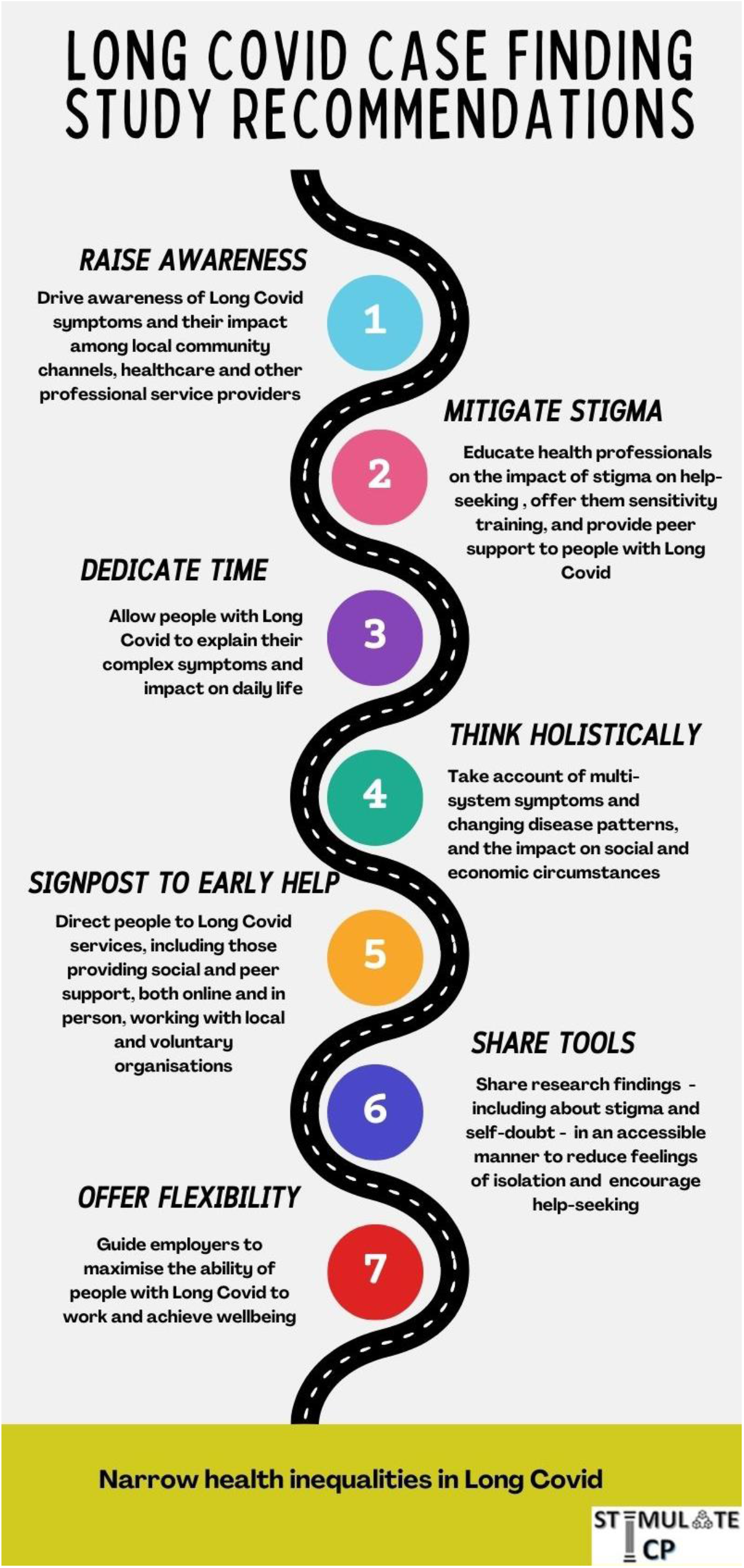
Long Covid active case finding study recommendations.

The co-produced nature of this study is a strength. Including the voices of people with lived experience of Long Covid has resulted in research and findings that are more relevant to others experiencing Long Covid symptoms. The case finding strategy used in this study offers one way to identify cases of probable Long Covid. Long Covid cannot be proven by one diagnostic test and is usually diagnosed based on symptoms. The eligibility and screening criteria used in this study are based on the WHO Clinical Case Definition of Post COVID-19 condition.^1^ This analysis did not include younger people (<29years), so we do not know if the experiences for people with Long Covid symptoms in their late teens or twenties are the same as for older age groups. Future research should aim to include the experiences of young adults.

We successfully employed a co-designed active case finding strategy to identify cases of probable Long Covid in three local communities in England. Our findings have highlighted the different, sometimes intersecting, barriers that can prevent people living with Long Covid from obtaining a diagnosis, treatment, or care for their symptoms. Despite Long Covid being a relatively new illness, it can be a stigmatising experience congruent with epistemic injustice. Action on raising awareness, mitigating stigma, listening to people with lived experience, and signposting to early help in the health and wider services are needed.

## Author Contributions

NAA, MP, MG, GA, AB and JH conceptualised the original idea and obtained funding for the original study as part of STIMULATE-ICP. NAA, DC, MP, JH, MAK, CH, CR, DB, MJ, EP, GJ, RC, SM, DW and MH designed the methodology for the active case finding study. FB, MF, NN and BC contributed to recruitment design and conduct. DC completed all data collection and led data analysis for the active case finding study. MR completed additional coding. DC, MR and NAA developed the thematic framework which forms the basis of this manuscript. DC, MR and NAA drafted the original manuscript. All authors read, reviewed, and approved the final manuscript.

## Acknowledgements

Our sincere thanks to all of those who contributed during the Community Advisory Board meetings. Thank you to our study partners and collaborators, including Donna Turnbull and Nasrin Rashid at Voluntary Action Camden, Allegra Lynch at Camden Carers, Bernd Halschka at Merton Connected, Colin Brummage and Tom McDonough at Camden Disability Action, and Hannah Stanier at Derby County Community Trust. We also want to extend our thanks to the participants who shared their experiences and contributed to this study.

This protocol is part of the STIMULATE-ICP study. An up-to-date version of STIMULATE-ICP Consortium members can be found on https://www.stimulate-icp.org/team. STIMULATE-ICP can be contacted at.

## Funding Statement

This work was supported by the National Institute for Health Research (NIHR) [STIMULATE-ICP grant number COV-LT2-0043] and NIHR Applied Research Collaboration (ARC) Wessex. The funders had no role in study design, data collection and analysis, decision to publish, or preparation of the manuscript.

MG is part funded by National Institute of Health and Care Research Applied Research Collaboration North West Coast (ARC NWC).

DW is supported by an Advanced Fellowship from the NIHR (NIHR300669).

The views and opinions expressed are those of the authors and are not necessarily those of the NIHR or the Department of Health and Social Care.

## Ethics

This study received ethical approval from the University of Southampton Faculty of Medicine Ethics Committee on 27^th^ July 2022 (reference number 72400).

## Data Availability Statement

The anonymised data that support the findings of this study can be made available on reasonable request from the corresponding authors.

## Open Access

For the purpose of open access, the author has applied a Creative Commons Attribution (CC BY 4.0) licence to any Author Accepted Manuscript version arising.

