## Supplementary material for "Barriers to healthcare access: findings from a co-produced Long Covid case-finding study"

**Supplementary materials**

Supplementary material A: Eligibility Criteria

Potential participants who engaged with the leaflet were encouraged to contact the research team to check eligibility. Study eligibility was based on confirmation of:

1. History of probable or confirmed COVID-19 infection at least three months before the time of recruitment
2. Prolonged symptoms* after COVID-19 lasting at least 2 months that cannot be explained by another condition
3. Symptoms’ impact on everyday functioning
4. No existing clinical diagnosis of Long Covid or uncertainty regarding diagnosis.

*Participants were included if they had three or more of the symptoms listed below:

Post-exertional symptom exacerbation (symptoms such as fatigue, difficulty thinking, pain recurring after remission or getting worse following exertion, either immediately or up to 72 hours post exertion), exhaustion, cognitive dysfunction (brain fog, memory problems, concentration problems, occasional confusion), breathlessness, headache, muscle aches, palpitations, chest tightness/pressure, chest pain, dizziness, sleep disturbance, joint pain, leg pain, pins and needles feeling, tinnitus, sore throat, cough, nasal symptoms, hoarse voice, abdominal pain, nausea, diarrhoea, chills, altered or loss of sense of smell, altered or loss of sense of taste, skin rash, loss of appetite and sneezing.

Those who fulfilled the eligibility criteria above were invited to take part in the study.

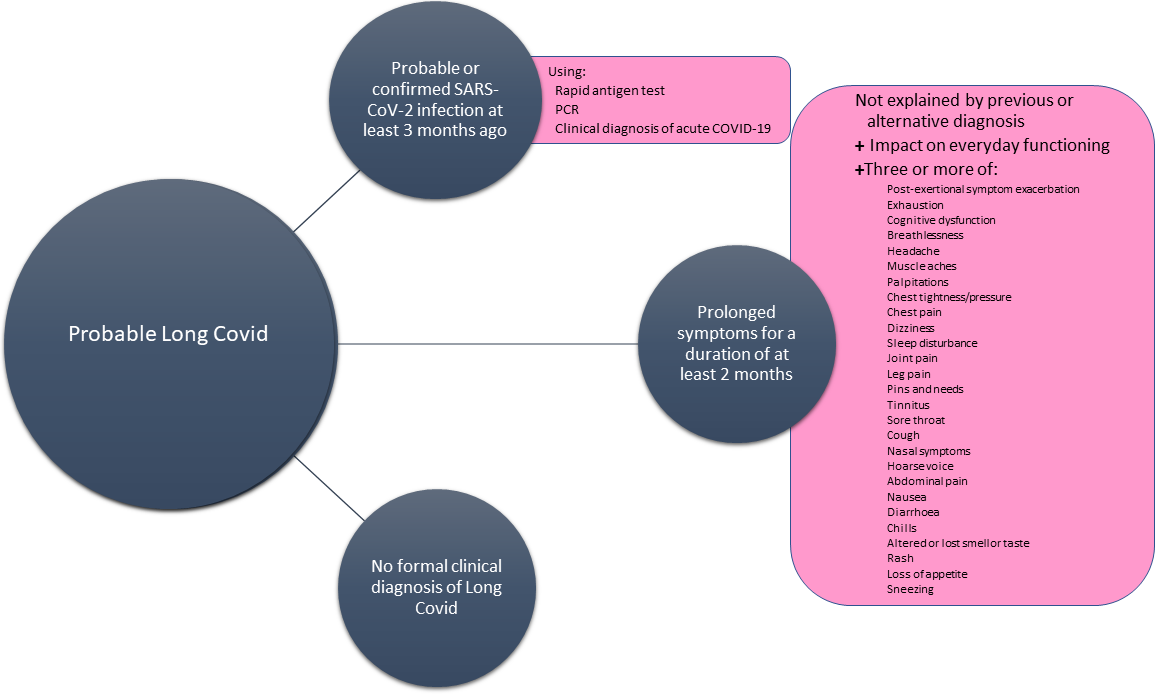

Image source: Alwan NA, Clutterbuck D, Pantelic M, Hayer J, Fisher L, Hishmeh L, et al. (2023) Long Covid active case finding study protocol: A co-produced community-based pilot within the STIMULATE-ICP study (Symptoms, Trajectory, Inequalities and Management: Understanding Long-COVID to Address and Transform Existing Integrated Care Pathways). PLoS ONE 18(7): e0284297. <https://doi.org/10.1371/journal.pone.0284297>

Supplementary material B: Screening Questions

**(To be administered by the researcher)**

**Researcher script (verbal consent to ask questions):**

*Thank you for contacting us about this study. Please can I ask where you heard about us?

………………………………………………………………………[Insert where the participant heard about the study here].*

*In order for us to check your eligibility for the study, please can I check that it is OK for us to ask you a few questions?*

Yes 🞏	No 🞏 - *If yes, continue to ask screening questions.*

1) a) Do you think you had Covid 3+ months ago?
    Yes 🞏	No 🞏 - *If yes continue to b)*

b) Was this suspected or confirmed Covid?
 Suspected 🞏	Confirmed 🞏 *If confirmed continue to c), suspected skip to 2)*

c) How was Covid confirmed?
 PCR test 🞏	Rapid antigen test (lateral flow) 🞏 Antibody test 🞏

2) Have you experienced prolonged symptoms after Covid infection?
   Yes 🞏	No 🞏 *– If yes, continue to question 3*
3) Do these symptoms restrict or limit your usual daily activities in any way (the usual activities you used to do before including work patterns, exercise and social activities)?

Yes 🞏	No 🞏 *– If yes, continue to question 4*

4) Have you been told by your GP that you have Long Covid?
   Yes 🞏	No 🞏 - *if yes, participant is not eligible to take part in the study*
5) a) Have you experienced any three of these symptoms for a duration of at least 2 months that are not explained by an existing health condition you already have – *If three or more are ticked from a) and b), participant is eligible to take part.*

- Exhaustion
- Brain fog
- Memory problems
- Concentration problems
- Occasional confusion
- Breathlessness
- Headache
- Muscle aches
- Palpitations
- Chest tightness/pressure
- Chest pain
- Dizziness
- Sleep disturbance
- Joint pain
- Leg pain
- Pins and needles feeling
- Tinnitus
- Sore throat
- Cough
- Nasal symptoms
- Hoarse voice
- Abdominal pain
- Nausea
- Diarrhoea
- Chills
- Altered or loss of sense of smell
- Altered or loss of sense of taste
- Skin rash
- Loss of appetite
- Sneezing

b) Do any of the symptoms described reoccur or get worse immediately or up to 72 hours following an increase in activity (PESE)?
   Yes 🞏	No 🞏

Those who fulfil the screening criteria above are assumed to have probable Long Covid and can be asked if they are interested in taking part in the study.

Supplementary material C: Symptoms

Participants suffered a range of symptoms consistent with Long Covid including exhaustion, brain fog, memory problems, concentration problems, occasional confusion, breathlessness, muscle aches, chest tightness/pressure, headache, dizziness, sleep disturbance, leg pain, tinnitus, cough, nasal symptoms, hoarse voice, nausea, diarrhoea, loss of appetite, sneezing, food allergies, altered sensations in the body, palpitations, skin rash and altered or lost sense of smell or taste. Some participants saw an exacerbation of existing conditions.

Participants reported experiencing symptoms for varying time periods, with some suffering from the acute COVID-19 infection in early 2020 and others in early 2023. Some had multiple infections. Most participants had this confirmed by polymerase chain reaction (PCR) or rapid lateral flow (LFT) tests, though five participants had suspected infections prior to the widespread availability of testing. Most participants were not hospitalised during the acute phase of their COVID-19 illness.
